# Evaluation synthesis analysis can be accelerated through text mining, searching, and highlighting: A case-study on data extraction from 631 UNICEF evaluation reports

**DOI:** 10.1101/2024.08.27.24312630

**Authors:** Lena Schmidt, Pauline Addis, Erica Mattellone, Hannah O’Keefe, Kamilla Nabiyeva, Uyen Kim Huynh, Nabamallika Dehingia, Dawn Craig, Fiona Campbell

## Abstract

**Background:** UNICEF works to protect children’s rights and improve their well-being by partnering with governments and communities through various programs. These include protection, education, health, water, sanitation, and emergency support. Publicly available evaluation reports^1^ provide insights on results, recommendations, and lessons learned.

**Objective:** This article describes semi-automated methods to synthesize UNICEF’s extensive evaluation report collection to assess UNICEF’s impact, identify achievements, challenges, and inform future strategy.

**Methods:** A semi-automated approach was used to extract data from 631 evaluation reports across 64 outcomes in UNICEF’s 2022-2025 Strategic Plan. Text pre-processing involved PDF-to-text extraction, section parsing, and neural network sentence mining. Data extraction was facilitated by SWIFT-Review using adjacency-search queries for outcome-based filtering.

**Results:** We reduced text volume by 92%, identified relevant sentences with high recall (0.93), and outcomes within evaluation texts with median precision 0.6 (reading 21 reports per outcome).

**Conclusions:** Semi-automation accelerates synthesis while maintaining scientific rigor and reproducibility.

**Plain Language summary:** UNICEF is a global organization that works to improve the lives of children by protecting their rights and helping them access essential services like education, health care, clean water, and protection from violence. UNICEF’s work is guided by a strategic plan and is evaluated through reports that describe their programs in detail. These reports, available to the public, contain valuable information on the results, recommendations, and lessons learned from UNICEF’s work around the world.

We would like to understand UNICEF’s impact and identify areas where they can improve. This can be done by summarizing the main achievements, challenges and opportunities for future strategies based on UNICEF’s evaluation reports. Because there are many reports, reviewing all of them manually would be too time-consuming, so a semi-automated process was used to speed up the work.

We developed a method that used computer tools to highlight key information from 631 evaluation reports. These reports covered 64 different outcomes based on the goals in UNICEF’s 2022-2025 Strategic Plan. We automatically turned text into machine-readable format, used artificial intelligence to identify important sentences, and then used advanced software to filter and find the most relevant information.

The semi-automated approach was effective, reducing the amount of text by 92%. It also identified relevant outcomes with high accuracy. This method helped us speed up the process while still maintaining the quality and reliability of our findings.

Overall, using automation helped save time and effort, making it possible to analyze large amounts of data efficiently while still ensuring the results were scientifically sound.

**Strengths and limitations of this study:** Systematic impact evaluation syntheses are a vital tool to critically evaluate and plan future work of organisations such as UNICEF; but they are often not feasible due to the size, structure, and amount of evaluation report documents.

To increase feasibility of analysis we describe semi-automated human-in-the-loop methods which were applied in a synthesis of 631 evaluations across 64 outcomes.

The proposed open-source code and methods made an evaluation synthesis feasible by reducing text and streamlining the identification of relevant reports for each outcome.

By making code open-source and adaptable we aim to encourage accelerated, yet transparent and reproducible results.

While the methods cannot produce 100% complete or correct results for each outcome, they present useful automation methods for researchers facing otherwise non-feasible evaluation syntheses tasks.

## Introduction

### Background

The United Nations Children’s Fund (UNICEF) is the United Nations agency dedicated to promoting and advocating for the protection of children’s rights, meeting their basic needs, and expanding their opportunities to reach their full potential. They achieve this working with governments, communities, and other partners via programmes that safeguard children from violence, provide access to quality education, ensure that children survive and thrive, provide access to water, sanitation and hygiene, and provide life-saving support in emergency contexts.

For this broad portfolio of programmes, UNICEF publish comprehensive evaluation reports to disseminate outcomes of their interventions and to accelerate results for children. These evaluation reports include a wealth of information on what has been achieved, enabling and hindering factors, recommendations, and lessons learned in the implementation of specific UNICEF projects. As such, the evaluations often are stand-alone, project- or programme-focussed reports, while some also represent efforts to synthesise existing evaluations. Between 2018 and 2023 UNICEF have published 875 such evaluation reports^2^.

Evaluation reports are commonly published by aid and non-profit organisations to communicate the methods and outcomes of projects. The International Initiative for Impact Evaluation (3ie), for example, maintains an online portal of more than 13,000 impact evaluations spanning sectors such as health, education, energy, or social protection^3^. The United Nations Development Programme (UNDP) maintains a database with more than 6000 evaluation reports and an in-built analysis platform^4^. Oxfam, a British group of non-governmental organizations, maintains a database of 88 impact evaluations^5^. UNICEF’s evaluation function covers thematic, humanitarian, real-time, country level and syntheses aimed at assessing impact, efficiency, and effectiveness of its programs and maintains a digital database of a broader range of evaluative reporting^6^ (see Figure 1). UNICEF evaluation reports are published in English, French, Spanish, or Portuguese and commonly include an executive summary, background, methodology, conclusions, lessons learned, recommendation for future programmes, references and multiple appendices with additional text, data, and tables in order to be transparent about the work that was carried out in the scope of each project. Therefore, they often exceed 150 pages of plain text, as commonly seen with reports from other governing bodies or leading organisations that conduct this scale of work. This complicates manual secondary analysis, due to the large amounts of unstructured, and potentially irrelevant, data. To increase the feasibility of analysing such a large, dataset in a timely, yet also reproducible manner, we developed a methodology that includes tagging and text-mining approaches to rapidly identify relevant data within the reports and expedite analysis.

**Figure 1:**
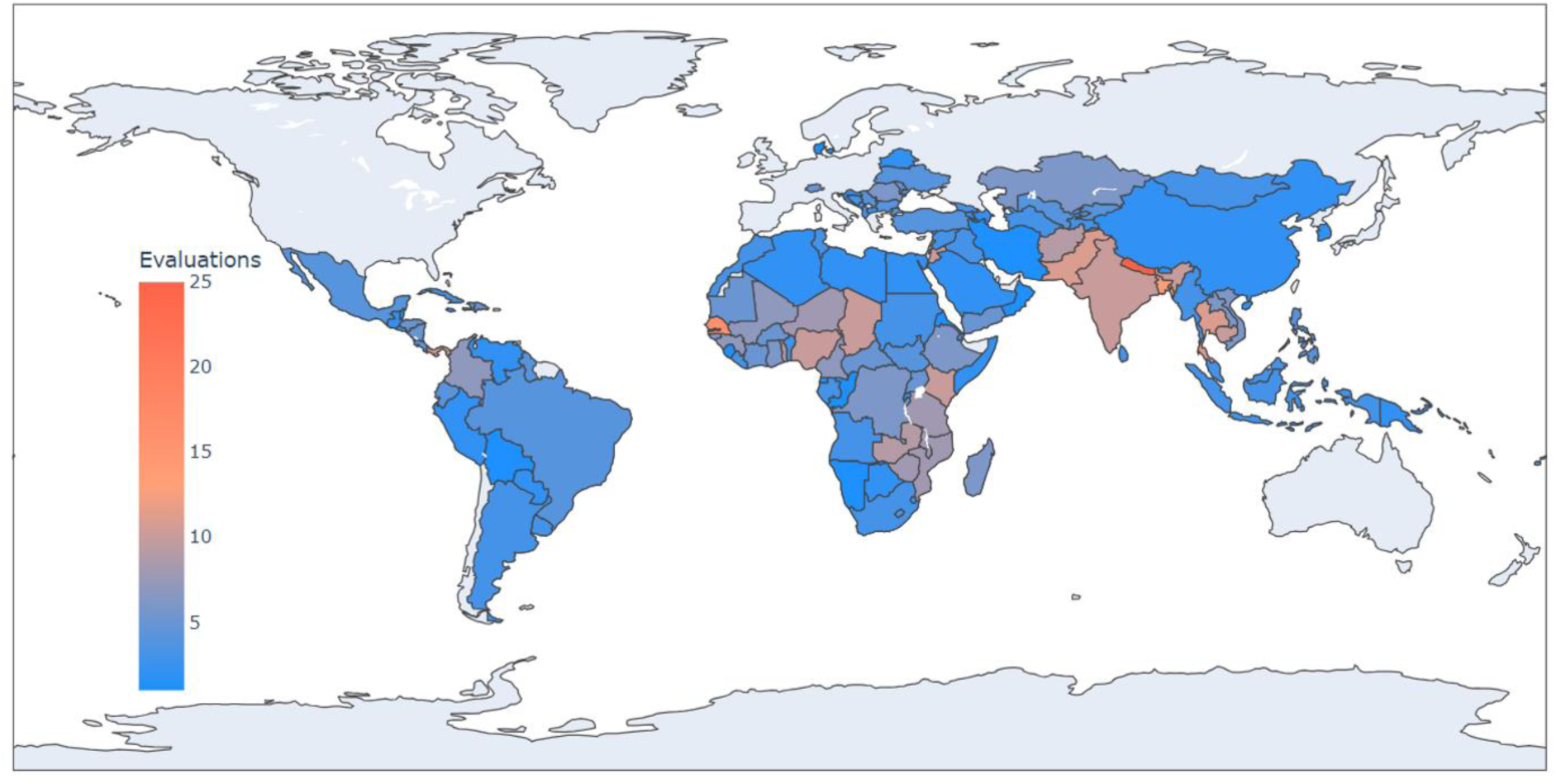
Number of UNICEF evaluation reports by country, visualising the global spread of programmes. We used information from UNICEF’s internal database to aggregate the number of evaluations published by each country office, published 2018-2022 with UNICEF evaluation quality-assessment system ratings above ‘Satisfactory’ level.

### Aim

The aim of this paper is to demonstrate how text mining was used in a human-in-the-loop system to make an evaluation synthesis feasible and sustainable, while keeping its scope as broad as possible.

We describe the process of synthesising information from UNICEF reports, with respect to 64 outcomes related to five goal areas formulated as part of the Strategic Plan for 2022-2025: ‘Every child survives and thrives’, ‘Every child learns’, ‘Every child is protected from violence and exploitation’, ‘Every child lives in a safe and clean environment’, and ‘Every child has access to inclusive social protection and lives free from poverty’ (UNICEF, 2022). Outcomes focus on specific problems within each goal area, for example school attendance rates, access to safe drinking water, or domestic violence. The present paper focuses on semi-automation methods that made this analysis feasible. All code and trained models are available via our GitHub (see Data Availability Statement). This paper primarily focuses on automation methods rather than the outcomes of the evaluation synthesis itself. For more information about the results of the evaluation synthesis, please visit the UNICEF website^7^.

### Sustainability and applicability

The automation and text-mining methods described in this paper were developed and tested as a case-study within UNICEF evaluation reports. These reports were created across more than 120 different country offices, they vary in length, and describe project across a diverse range of topics, such as sanitary infrastructure, cash transfers, vaccinations, or IT infrastructure. Reports from other organisations such as 3ie or Oxfam similarly are likely to differ in length, structure, and content, hence we have aimed to create generalisable methods that are easily adaptable. Additionally, by making the code and text-mining methods open-source, and by using third-party software that is free to use and available to the general public, we have tried to ensure that our methods are available to other researchers who wish to apply them to synthesise similar report datasets.

### Methodology

The following section introduces the dataset and automation pipeline in our case-study. We divided the process into several steps. For each step we provide open-source Python code on GitHub, developed as part of the case-study, to encourage adaptations and use in different research contexts. Figure 2 displays the methodology and number of reports included in each stage, in the form of a flowchart.

**Figure 2:**
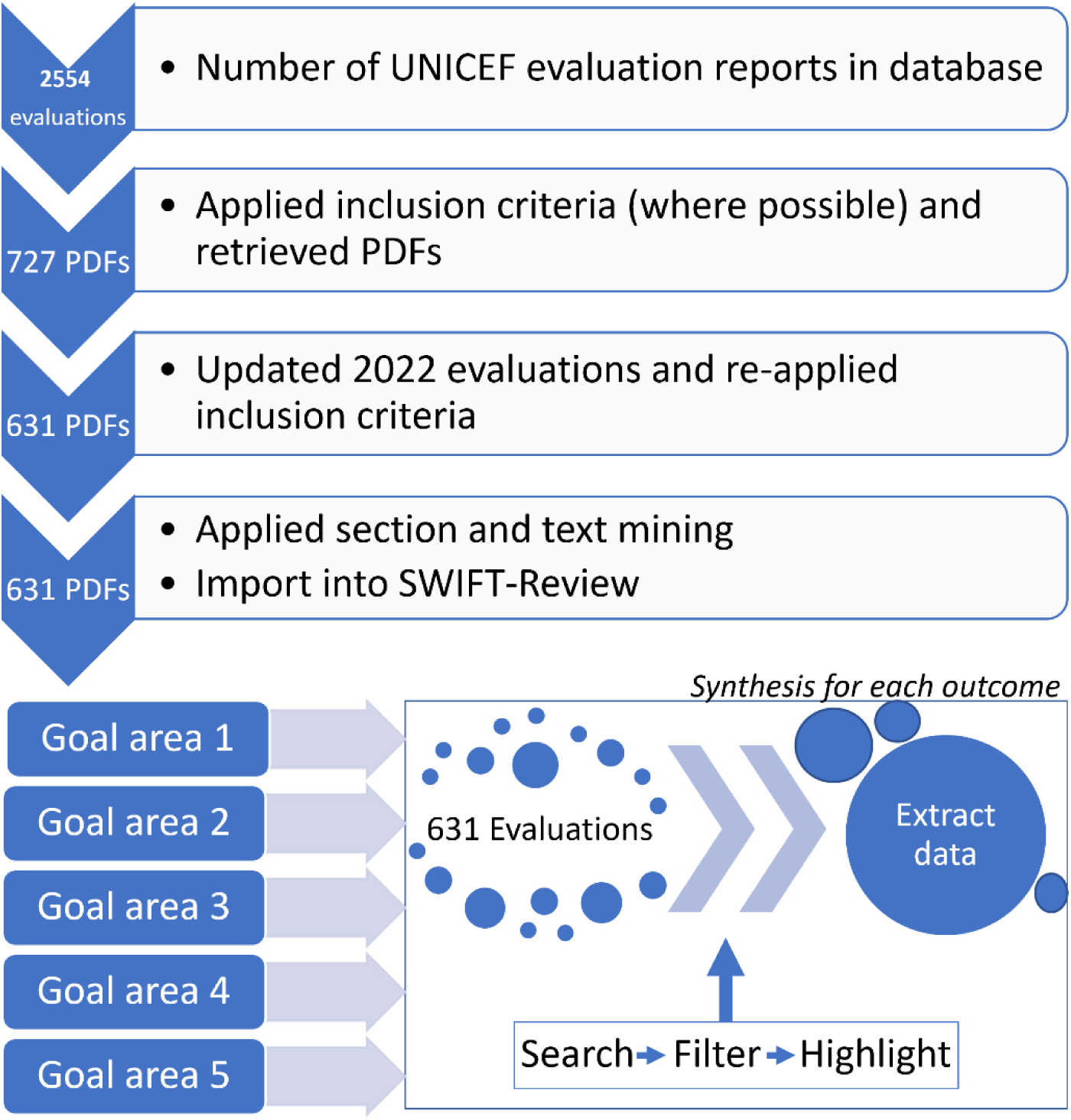
Methodology flowchart, displaying the number of evaluation reports included in each stage of the analysis. We used information on publication year and report quality ratings within UNICEF’s internal evaluation database to apply inclusion criteria. Identification of relevant text sections, sentence mining, as well as the process of identifying relevant reports for each goal area and outcome supported by our scripts and third-party software are described in more detail in the methodology section of this article.

**Figure 3:**
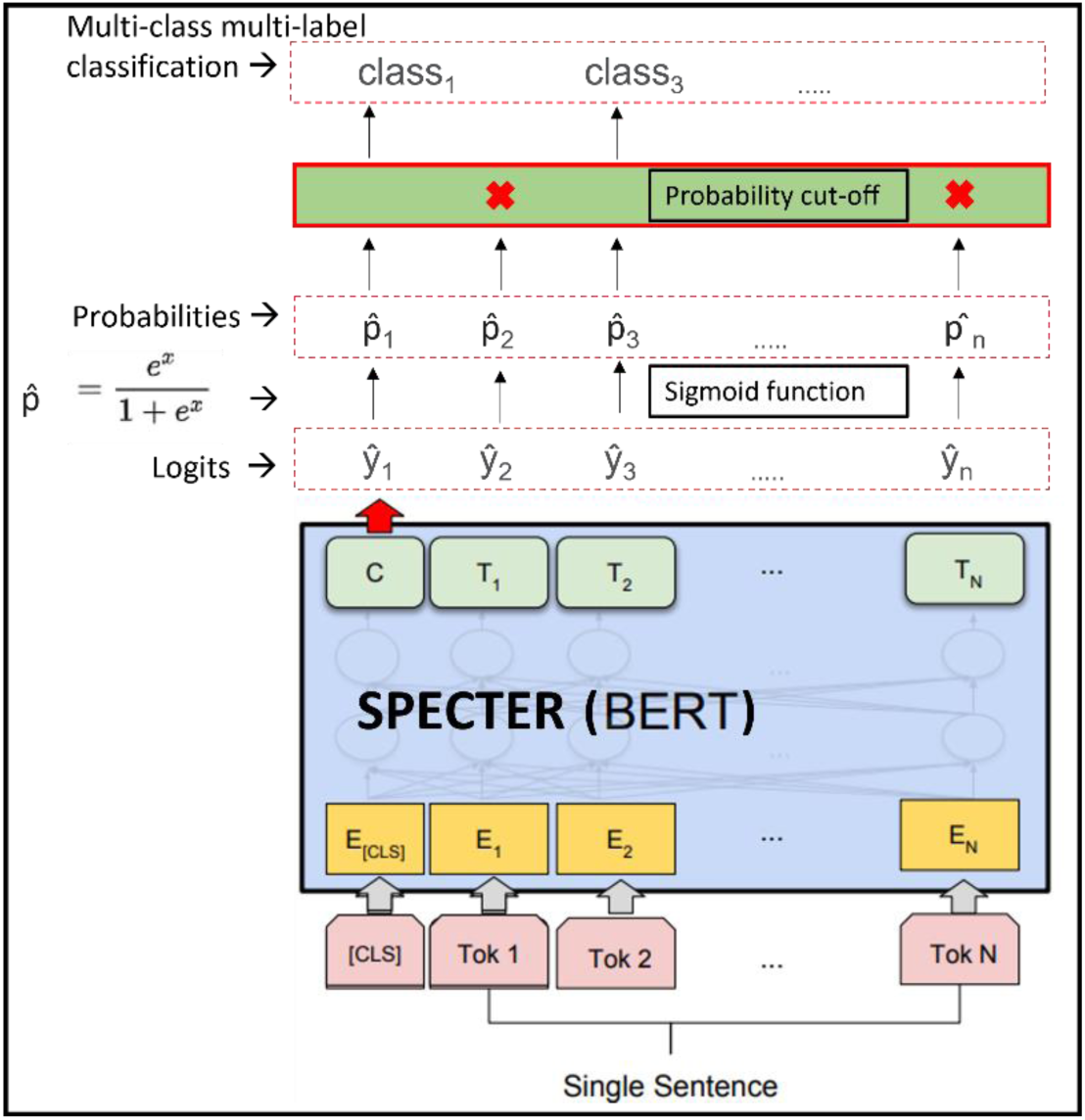
Neural network architecture. Plain text is tokenised into sentences and then embedded using the pretrained SPECTER network. We train, evaluate, and then use this neural network on evaluation report full text data to obtain probabilities across all target classes for each input sentence. Initially, the neural network output is a vector of logits, which we transform to probabilities using the sigmoid function. On the resulting probability vectors we apply a cut-off value of 0.2 for classification, thus creating a multi-class multi-label prediction scenario. Figure adapted from Devlin et al. (2018).

### Dataset

PDFs of 727 reports were obtained with the help of UNICEF, from their public evaluation reports database^8^. We included all reports that fulfilled the following inclusion criteria:

1. Published between 2018-2022
2. Reports previously rated as ‘Satisfactory’ or better, using UNICEF’s *evaluation quality- assessment system*; GEROS ^9^

After updating the data with the latest 2022 evaluations and re-applying the inclusion criteria, 631 evaluation reports remained within the final dataset.

### Automation methodology

The automation steps are as follows:

1. Bulk PDF-to-text conversion
2. Identification of relevant sections of text using rule-based methods and mining sentences using a neural network
3. Use of SWIFT-Review (Howard et al., 2016) to filter reports and highlight text for data extraction. Additionally, we provide a script to support the development of comprehensive adjacency-based search strategies
4. Automatic translation of non-English text

#### 1. Bulk PDF-to-text conversion

We used the Python package *pypdf*^10^ to convert report PDFs into text. First, the script accesses a folder where PDFs are saved. It then iterates through all files and attempts to read them as PDFs. Next, it accesses the PDFs page-by-page, extracts text and saves it as plain text file in a different folder. In rare cases there may be PDF processing errors, to mitigate this limitation our script records the names of affected files.

#### 2. Identification of relevant sections of text using rule-based and neural methods

##### Rule-based methods

We tested automatically extracting executive summary, lessons, and recommendation sections as these were deemed to be most likely to include relevant information. Initially, a rule-based approach was tested: matching text and extract sections by identifying words that would appear in a section heading, including synonyms and translations into French, Spanish, and Portuguese. For example, a case-insensitive regular expression search for ’(lessons)|(lições)|(leçon)|(lecciones)|(good practice)’ in the vicinity of a numbered item and carriage returns might indicate a section header for ‘Lessons Learned’. However, it is very challenging to identify the end of a section, as some but not all reports include subsections or multiple paragraphs. Due to this problem, and additional variability in section header names, the resulting text for lessons and recommendation sections was not of sufficient quality.

Executive summaries, however, were identified automatically using the case-insensitive regular expression ’(executive summary)|(RESUMEN EJECUTIVO)|(RÉSUMÉ EXECUTIF)|(Résumé exécutif)|(SOMMAIRE EXECUTIF)|(Resume executif)’. Due to this section’s reliable placing at the beginning of a report and a limitation to 12 pages of text, we were more confident to use automatically extracted text from this section for the analysis. We were unable to identify 22% of the executive summaries automatically due to variations in wording or structure and quality of the PDF documents. These executive summary sections were extracted and added to the dataset manually.

##### More advanced text-mining from full texts to raise data quality

One researcher spent around three hours processing ten randomly-chosen evaluations and identifying relevant sentences describing ‘Enablers’ (N=55), ‘Barriers’(N=57), ‘Lessons Learned’(N=151), ‘Recommendations’(N=190), and ‘Background’(N=422) from their respective sections within the report’s full text. The first four categories include the target information that is useful for this project, while the ‘Background’ class includes a mixture of undesirable text, such as table of contents, abbreviations, introduction, or methods sentences.

Using the sentences for each of these categories, a neural network based on the transformer architecture (Devlin et al., 2018) and a previously published model called ‘SPECTER’ (Cohan et al., 2020) was trained to identify relevant sentences of each category. We also tested the performance of other models such as BERT^11^, SCIBERT^12^, BERT-mini^13^, and XLNet^14^ (see code on GitHub) but SPECTER showed the strongest results. A potential reason for this is that it was pre-trained on scientific data using a training objective that promotes the identification of semantically similar data; which is useful for downstream tasks that aim to classify related text (Cohan et al., 2020). A random split of 60% of the data were used for training, and the rest for evaluation. This dataset had limitations: being very small, labelled rapidly, and containing classes with senses that are ambiguous and overlap. However, this makes it an excellent test case for applying text mining methods on future projects analysing large amounts of unstructured grey literature, to maximise research outputs with respect to very tight timeframes and low resources available for analysis.

As this model was trained exclusively on English data, 162 non-English evaluations that were previously identified by UNICEF to be either French (n=96), Spanish (n=63) or Portuguese (n=3) were translated using the freely available Google Translate API and the *googletrans* python package^15^. All documents were then split into sentences. Pre-processing methods such as removing encoding errors, page breaks, and sentences shorter than 20 characters were applied to reduce noise in the dataset. Every sentence passed through the classifier and sigmoid layer and received a prediction of the likelihood of belonging to each class, thus creating a multi-class multi-label prediction scenario (see Appendix 2). When we applied the model to the full dataset, for each evaluation document the 30 sentences with the highest probability scores were chosen.

To increase sensitivity, for each category some additional sentences were chosen, for example for ‘Lessons learned’, the filters ‘need to’, ‘may’, ‘lesson’ were applied to all sentences and the 30 most likely sentences containing each term were also added to the final dataset if they were not already contained within the model predictions. The ordering was based on the model’s predicted probabilities for this class, highest first.

#### 3. Semi-automating data extraction with SWIFT-Review

After the text-mining step, each original evaluation report was reduced to a plain text file including an abstract-length short description provided by UNICEF, the report’s 12-page executive summary, and mined sentences. We created a RIS file that contained the content of these text files within the abstract field, as well as metadata for each report in the form of keywords. This allowed us to import the data into third-party software for further analysis.

To facilitate the full analysis of the final 631 evaluation reports for each of the outcomes, we imported the data into a SWIFT-Review project. SWIFT-Review is a freely available text-mining workbench that facilitates searching, keyword highlighting, and topic modelling (Howard et al., 2016).

We created comprehensive tagging strategies for each outcome, to rapidly identify relevant information within each report. SWIFT-Review includes advanced search functionalities to tag or highlight text in fields such as title, abstract (which in our case included all text data described earlier), keywords, and pre-processed versions of the text such as stemmed versions. It also allows large Boolean searches, combining clusters of terms with ‘AND’, ‘OR’, and ‘NOT’ operators, wildcards, and adjacency searches to find documents containing target terms within a distance of N words.

Due to the complexity of our outcomes of interest, and the length of the documents, we opted against using simple Boolean searches to tag documents. For example for outcome 1.8 “*Percentage of surviving infants who received (a) first dose and (b) three doses of diphtheria, tetanus and pertussis (DTP) vaccine (WHO)*” using search terms *(’diphtheria’ OR ’tetanus’ OR ’lockjaw’ OR ’pertussis’ OR ’whooping cough’) AND (’vaccine’ OR ’vaccinated’ OR ’immunisation’ OR ’immunization’ OR ’jab’ OR ’injection’)* would have meant that every document using these terms anywhere would have been selected; even if they were not in direct context. To increase precision of our results we instead set the search up to use adjacency searching, thus combining every word in the first Boolean arm with each word in the second arm and allowing a default of up to 5 words between target terms.

For each outcome, between one to three search arms were devised to search, filter, and highlight data for extraction. A data scientist and an information specialist created the initial versions, and a senior reviewer responsible for the final data extraction reviewed and extended them to maximise sensitivity of the results. To save time, a python script was used to combine the terms from each arm of the search into an adjacency search query on SWIFT-Review’s “tiab_stemmed” field. This field includes a pre-processed version of the text where only word stems are used to match text, thus reducing the need for wildcards. After generating and running the searches on our set of 631 reports we adjusted them as needed, either by decreasing the default 5 words for the adjacency to decrease the number of hits, or by skimming results from outcomes with a high number of hits to remove terms that appear to retrieve irrelevant information.

This led to search strategies such as *’tiab_stemmed:“adolescents school dropped out”∼5’* to filter evaluations. That search, in practice, filters all evaluation where the words “adolescents school dropped out” and their grammatical variations appear within the proximity of 5 words of each other, eg. in “The scholarship programs and monitoring of children and **adolescents** who have **dropped out** of **school**”.

Figure 4 shows how the search for outcome 1.8 was applied to the 631 documents in SWIFT-Review. The search itself was pasted into the query field on the top left corner of the screen. After executing the search, SWIFT-Review matched 12 evaluation documents that are selectable on the bottom half of the screen. After selecting a document, SWIFT displays the document text in the top right corner, with yellow highlights applied on the words that matched the search. Due to overlaps and similar content between outcomes, we grouped some together for screening and data extraction, reducing the total number of separate outcomes to 34 (data shown in Appendix 1). For example, outcomes 1.8 and 1.9 were combined (1.8: ‘Percentage of surviving infants who received (a) first dose and (b) three doses of diphtheria, tetanus and pertussis (DTP) vaccine (WHO)’, and 1.9: ‘Percentage of surviving infants who received first dose of the measles-containing vaccine’). This allowed us to rapidly filter and skim relevant evaluations for each of the outcomes while avoiding duplication of effort.

**Figure 4:**
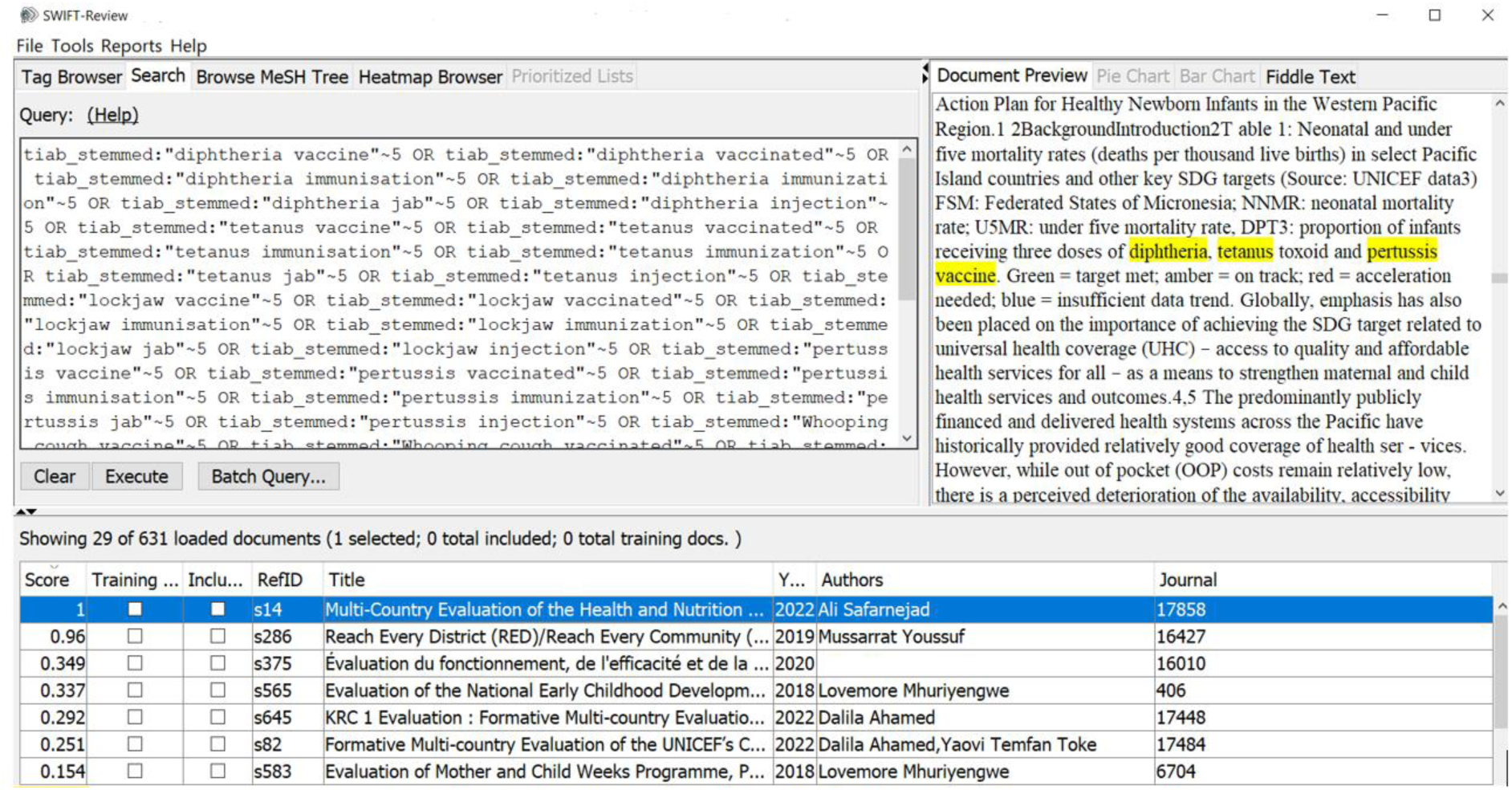
Example output filtering and highlighting 29/631 evaluations for outcome 1.8 and 1.9. The top left window shows our systematic Boolean search with adjacency queries for outcome 1.8 + 1.9. Those outcomes were merged due to their similar topic. The top right shows the original evaluation report text, with highlights where the search query matched. On the bottom, the user can select every report that produced at least one text match to the query, with reports that contain multiple matches automatically sorted to the top.

#### 4. Dealing with multiple languages and automating translation

UNICEF’s metadata for each report indicated the presence of French (n=96), Spanish (n=63) or Portuguese (n=3) evaluations. The neural network supporting the identification of ‘Lessons’ and ‘Recommendation’ sentences was trained using English text. Therefore, we used the GoogleTranslate API via the python package googletrans^16^ to translate these documents into English before predicting sentences.

In the SWIFT-Review-supported part of the project we trialled two approaches to handle non-English data.

Dataset 1: We used googletrans to translate any non-English text wherever this was possible. This included the abstract and executive summary sections, as well as the previously translated mined lessons and recommendation sentences from the full text.

Dataset 2: We used the original language abstracts and executive summaries. For the additional mining of lessons/recommendation sentences we had to use translated English text because the neural network was only trained on English data.

UNICEF then provided translations for all our terms in the goal area (GA) 2 outcomes searches. We re-created the search strategies for these terms in French, Spanish, and Portuguese to compare results for filtering.

Steps:

1. Run GA 2 searches in English on Dataset 1 (English translations)
2. Run GA 2 searches in French/Spanish/Portuguese on Dataset 2 (original French/Spanish/Portuguese)
3. Check if the translated searches from step 2 bring up any unique new hits that would be missed had we only used Dataset 1
4. If yes - consider translating terms for all GAs moving forward
5. If no - consider working with English/translated data only

We manually reviewed all evaluations that were filtered by the original language searches, but ’missed’ by the translated search. Data is shown in Appendix 2. Errors mostly occurred due to American/British English variations and ambiguous words such as the french ‘cours’ which can mean ‘lesson’ or ‘during’ and thus identify false-positive documents. Given the reasonable results of the error analysis we decided to use only automatically translated English text going forward. This led to time savings by avoiding the re-creation of all searches in three more languages and meant that no additional researchers were needed to perform data extraction from the majority of non-English language texts. To avoid missing data, we adapted the existing searches to include American and British spelling variations.

## Results

After automated PDF-to-text extraction, we automatically identified 88% of the ‘Executive Summary’ sections that contain most of the data of interest for the evaluation synthesis. However, there may be additional information throughout the reports, which is why we trained a sentence-classification model to retrieve information from the remaining report full texts, as ‘bonus’ information.

The evaluation results for this sentence classifier are shown in Table Table 11, showing an acceptable recall level of 0.93 for our ‘Lessons Learned’ and ‘Recommendation’ classes of interest. These scores were calculated using the scikit learn classification report function.^17^ We report scores for each class separately. For ‘Enablers’ and ‘Barriers’ it was challenging to identify training and evaluation data of sufficient quality. Evaluation reports often contain dedicated sections for Lessons and Recommendations, while Enablers and Barriers are described throughout the reports.

**Table 1:**
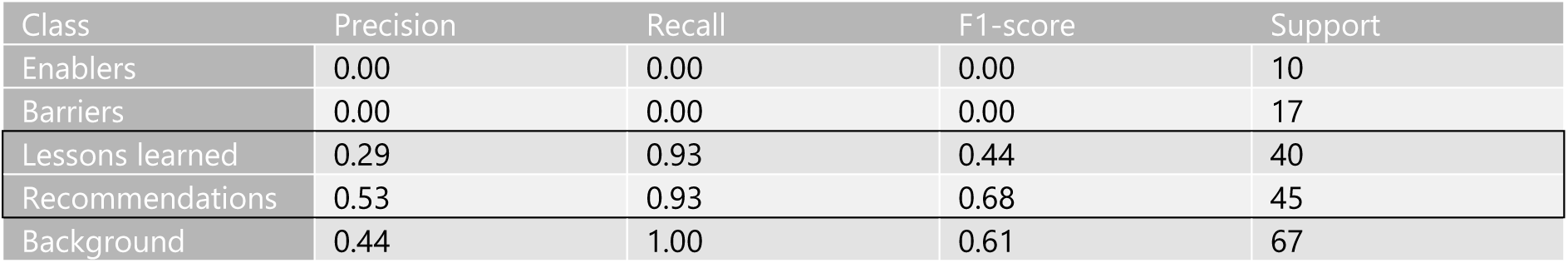
Evaluation of the text-mining model, assigning positive labels at a probability threshold of 0.2. High recall (i.e. sensitivity) shows that the model identified 93% of the relevant sentences for lessons and recommendations on the independent test set. We used the classifier only to predict these two classes on all data from 631 evaluation reports. The first three columns show quantitative evaluation results. The last column, ‘Support’, indicates the number of labelled evaluation samples from the held-out dataset that was used to calculate results.

| Class | Precision | Recall | F1-score | Support |
| --- | --- | --- | --- | --- |
| Enablers | 0.00 | 0.00 | 0.00 | 10 |
| Barriers | 0.00 | 0.00 | 0.00 | 17 |
| Lessons learned | 0.29 | 0.93 | 0.44 | 40 |
| Recommendations | 0.53 | 0.93 | 0.68 | 45 |
| Background | 0.44 | 1.00 | 0.61 | 67 |

Additionally, the latter two classes are challenging to define, highly ambiguous, may overlap with ‘Lessons’ and ‘Recommendations’ and when negated an Enabler can become a Barrier and vice versa. In the absence of high-quality data, the model did not adjust its weights sufficiently during the training process to classify sentences above our chosen threshold of 0.2. We did, however, decide to report results in full and to retain all classes so that the model could make use of all the limited training data that was available.

We then applied the trained neural network to the texts of all 631 evaluations. For each sentence, we transformed the neural network’s output into a vector that contained the probabilities of this sentence belonging to each of the five classes. We selected the top 30 sentences for the well-performing ‘Lessons Learned’ and ‘Recommendation’ classes each. Additionally, we filtered all sentences from each report using a keyword list for those two classes. This would lead to a theoretical maximum of 120 sentences for each report when considering both lessons and recommendation classes. However, the final number was usually lower than 120, for example obtaining 69 out of 2602 sentences for report 405. In the following, we report mean and standard deviation (SD) number of sentences that were retained by our algorithm. The reports included an average of 1662 sentences (SD 875), an average of 69 sentences (SD 21) were retained as ‘Lesson Learned’. For ‘Recommendations’, the average was 79 sentences (SD 22).

A merged version of lessons and recommendations was created, leading to an average of 143 unique sentences per report (SD 39). This text-mining step therefore reduced the total text volume down to 8% of its original size, when taking the average of 1662 sentences in a full report as baseline. We appended these mined sentences to the executive summaries for each report.

Using vocabulary provided by UNICEF and selected outcomes from each goal area and research output of interest, we created comprehensive search strategies to filter and highlight documents within SWIFT-Review. This included proximity searches to expand the result set, as well as structured data imported from a UNICEF database to tag reports by year or quality ratings, among other variables. We first applied this methodology to goal area 2 outcomes in a pilot. Given the positive results, we decided to move forward with this approach in the full analysis of all outcomes. We recorded which evaluation reports were screened and included in the analysis of each outcome and visualised the country of their respective UNICEF evaluation office in Figure 5. This indicates that our synthesis effort managed to include an even spread of reports on a global scale, despite using text-mining and filtering to accelerate data extraction. There may still be potential biases in the overall dataset, due to differences in resource availability and the thematic foci of programmes conducted by UNICEF across regions and goal areas. For a more detailed breakdown of evaluations included for each goal area, see visualisations in Appendix 4.

**Figure 5:**
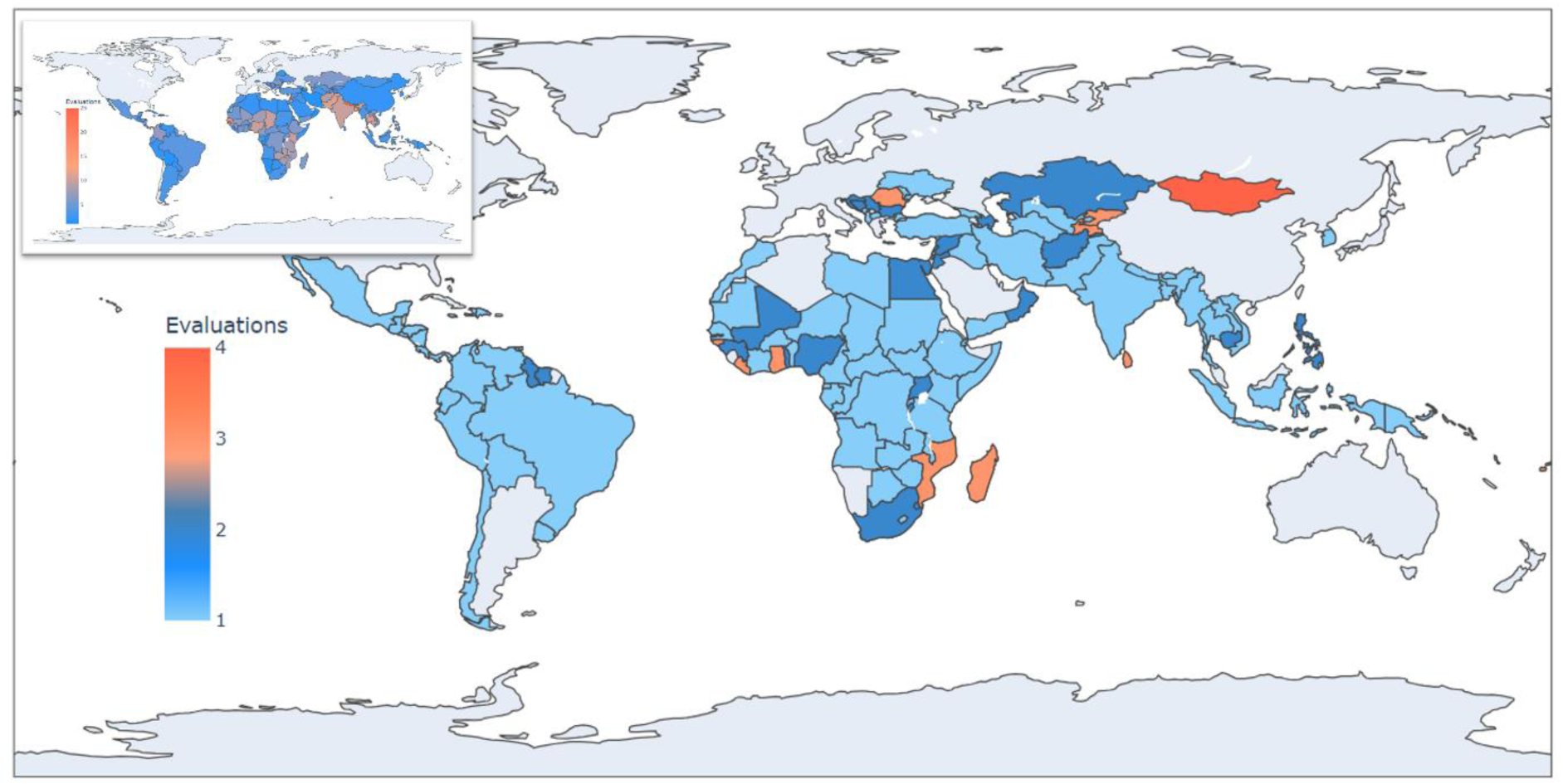
Global spread of evaluations filtered via SWIFT-Review and included in the synthesis of all 64 outcomes in five goal areas. For reference, a thumbnail of Figure 1 from this article is shown in the top-left corner, visualising the actual global spread of all 631 included evaluation reports.

Automatic searching and highlighting text from report summary sections for each goal area helped to reduce the overall workload of the analysis. This approach enabled researchers to quickly skim highlighted text of the most likely relevant evaluations, and to discard irrelevant evaluations. For evaluations with relevant highlights this approach allowed us to screen information more quickly, taking approximately 10-20 minutes per evaluation report as measured during a pilot phase for goal area 2 outcomes, although this could be up to 30 minutes when the original PDF had to be consulted. Among all outcomes, the precision of the filtering and tagging in SWIFT-Review was 0.52, having screened a total of 730 evaluations with 386 true positive evaluations that were included in the analysis.

On a per-outcome basis, precision values had a broad range between 0.08-1, with a median of 0.6 (full data shown in Appendix 1). The outlier precision score of 0.08 (seen in Figure 6a) was caused by outcome 2.5, where our workflow identified 12 reports to screen, of which only one was included during manual review. This outcome was from goal area 2, ‘*Percentage of countries in which the percentage of national education expenditure reaching the most marginalized is above 15 per cent’*. This is one of the more complex, less specific outcomes with expected lower coverage within the dataset. Therefore, we created a sensitive search with 27 term combinations to maximise chances of picking up relevant evaluation reports. Despite the low precision value of 0.08 the result was deemed acceptable because in terms of raw numbers, only 12 reports needed to be assessed, and one was identified as relevant. Outliers in terms of reports needing to be screened were for two outcomes with >70 results (seen in Figure 6b). These outcomes were related to the important topics of drinking water/WASH access and primary school enrolment rates, where large amounts of relevant evidence were expected and therefore we were not surprised by these outliers either.

**Figure 6:**
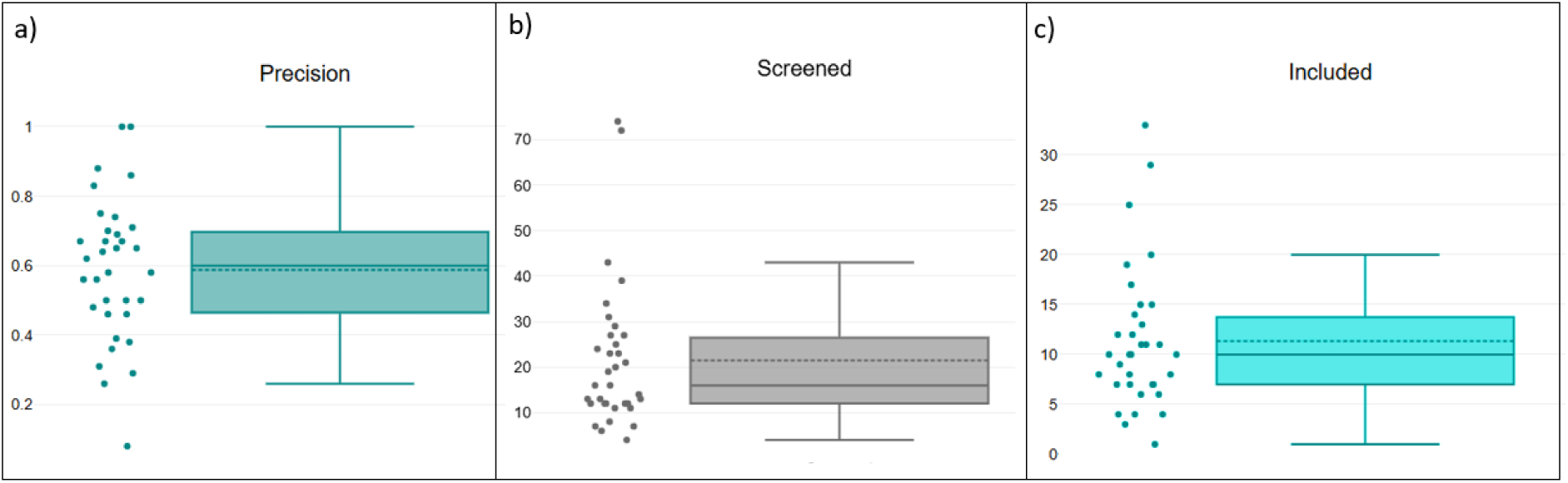
Results of our semi-automated approach, shown as box plots. The dashed line shows mean, and the solid line the median value. Dots represent outcomes and their associated values on each plot. Plot a) shows the precision values attained for each outcome. We defined True Positive reports as these who were picked up by our combined text mining and search methodology and assessed as relevant during manual review. False Positives were those reports picked up and later classified as irrelevant. Plot b) shows how many reports were screened for each single outcome. Plot c) shows how many reports were included for each outcome. In all plots, both very rarely occurring outcomes and very frequent ones may be outliers. However, this is to be expected when analysing such heterogeneous datasets.

Overall, our workflow reduced the number of evaluation reports to read for each outcome down to an average of 21 (seen in Figure 6b). In our dataset of 631 documents, this means that only around 3% of the evaluation reports were skimmed with keyword-highlighting assistance to assess each endpoint. Figure 6 provides some context on the relationship between precision and the total number of reports that were assessed and deemed relevant. More data, including full numerical data linked to outcome names, is provided in Appendix 1.

## Discussion

### Related literature and analysis tools

There are limited examples of tools or semi-automated methods to evaluate bodies of evidence such as these. We identified one tool developed by the United Nations Development Programme (UNDP), which is one of the six United Nations programmes.^18^ The UNDP maintains a database with more than 6000 evaluation reports. This database contains an in-built analysis platform that shares key traits with our proposed methods. Released in 2022, Artificial Intelligence for Development Analytics (AIDA)^19^ makes the plain text of UNDP evaluation reports accessible down to paragraph-level and provides features such as automatic summarisation. A comparison of current features is shown in Table 2.

**Table 2:**
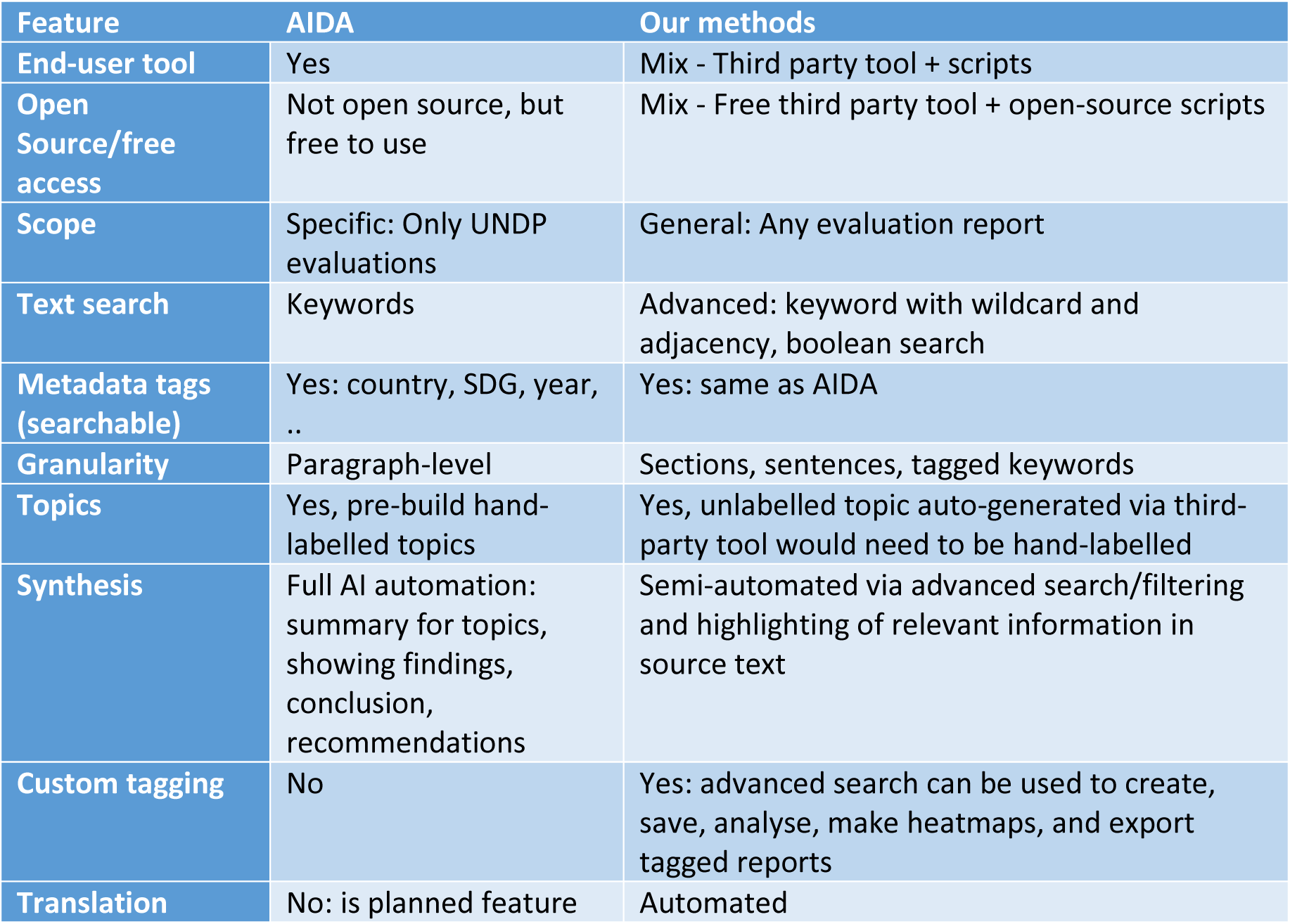
AIDA feature comparison with our proposed methodology

### Strengths

AIDA contains evaluations tagged with Sustainable Development Goals and other thematic keywords. It lets the user filter results by country or publication year, among other options. As such, AIDA serves the same purpose as our proposed methodology, by supporting researchers to filter, summarise, and visualise information. It surpasses functionality of our methods by providing fully automated synthesis outputs, which are useful for extremely rapid analyses. Another clear strength of AIDA is that it is a single, deployed end-user tool while our proposed methodology is a mix of end-user tools and open-source methods.

### Weaknesses

Key differences to our methodology are that AIDA is integrated into the UNDP infrastructure, not open-source, and thus not available to analyse other evaluation report datasets. While it supports keyword searching, it does not support complex search queries or highlighting of results in source text. Exportable results are AI-generated summaries and paragraphs with source text. We were unable to identify published information beyond a blog post^20^ and website. Questions remain about the underlying AI methods and how they were validated (ie. how well they work? How complete is the output? And if LLMs (Large Language models) are used to create summaries, are these affected by hallucination?). We appreciate that a full systematic validation of automated synthesis quality across 6000 evaluation reports for all (or even a few) SDGs is not feasible. However, smaller case-studies looking at specific SDGs in a rigorous systematic way might be able to assess if a human would have come to the same conclusion as AIDA, or if any of the two approaches (human vs. AIDA) would have missed important information.

### Usage in evaluation synthesis

We identified one UN-wide synthesis of SDG 5 (gender equality) evaluations that used AIDA and UNDP data; but also searched 53 further UN agency databases (Tanner et al., 2024). For UNDP data, AIDA was used to retrieve reports and was described as “powerful method for identifying UNDP evaluations related to specific SDG targets” (Tanner et al., 2024). However, as part of their more rigorous evaluation synthesis, AIDA output was used only to facilitate information retrieval and screening evaluations for inclusion. Afterwards, a random sampling approach was utilised to make the synthesis feasible, rather than relying on AI summaries (Tanner et al., 2024).

For search, our proposed methodology has similar features to AIDA’s, but is applicable beyond UNDP. Our method of systematic targeted search and highlighting of relevant source text for specific SDG sub-goal syntheses may offer faster, flexible, and more complete access to ‘rare’ outcomes that are not mentioned across many reports. The random sampling method used by Tanner et al. (2024) has a different strength: it is less likely to be affected by bias - and therefore more rigorous for mapping evidence of frequently mentioned goal areas and programme topics.

We are currently unaware of any other tools in this space, but equally are aware that internally facing tools are often difficult to find.

### Applicability of methods to datasets from other organisations

The flexibility and scalability of our methods is a strength, especially when compared with the AIDA tool that is currently confined to a single evaluation database. The PDF-extraction and sentence classification methods can be run locally or on the free tier of Google Colab (for GPU access). All methods can be applied to large corpora. Researchers from other organisations would be required to substitute training data for the sentence classification model with their own relevant data and re- train it, if they choose to use this model. However, if they prefer to use abstracts, selected summary sections, or other text available to them then they may skip this step. The length and complexity of Boolean search queries within SWIFT-Review to highlight outcome data is not limited. If others are interested in synthesising outcomes related to UNICEF’s goal areas, then they can re-use our queries from Appendix 1. If they require different queries, then our scripts to combine multiple search terms for adjacency queries can assist in creating large and comprehensive queries. The amount and length of records within a project is only dependent on the computing power available to the SWIFT desktop application (in an unrelated project we have seen up to 300.000 documents being searched, tagged, and highlighted, using a laptop with Intel i9 processor and 63GB physical memory).

When applying this methodology to evaluation report synthesis of other organisation’s data, we would like to highlight the following potential sources for error, and recommendations to mitigate them:

- Missing data due to PDF-to-text errors: We recommend using well-formatted and non-corrupted PDF files. Our script provides terminal output if text extraction fails, if this is the case then plain text needs to be copied from the PDF manually.
- Missing data from the sentence-classification models: We recommend a hybrid approach (as demonstrated in this paper) of using readily available summary data such as abstracts, short summaries, or executive summaries from evaluation reports in conjunction with the text mining model. This ensures that relevant information will be available if sentence classification is incomplete.
- Missing reports due to inadequate/too precise search queries: We recommend to involve an information specialist in the process of creating queries to retrieve reports for outcomes of interest. Generally, sensitivity (ie. retrieving as many relevant records as possible) should be preferred over precision. However, trade-offs may be required depending on the scale of project and available human resources.

## Strengths and limitations

The main strength of the rapid evaluation synthesis methods presented in this paper is that they present a resource-efficient way of analysing an extremely broad body of evidence. Due to variations in natural language and complex report structures, we are not claiming that the results of data extraction for each outcome are 100% complete. However, by using automatic translations, a human-in-the-loop system and systematic Boolean searches to filter the data, we add a degree of methodological rigour; promoting transparency and reproducibility within the process that isn’t given when using completely automated LLM-based systems. The search strategies for each outcome are shared in Appendix 1, and the SWIFT-Review project, which can be opened with the free SWIFT-Review desktop application^21^, is in Appendix 3. The programming code and automations we developed ourselves are available in a GitHub repository (See Data Availability Statement).

The main weakness on the methodological side, as mentioned above, is the chance of missing information during the data extraction process. This can happen at two distinct steps in the workflow. First, during the initial literature curation when only the most likely relevant information is retained. Secondly, due to natural language variations, information can also be missed when applying the searches and filtering in SWIFT-Review. Here, the project team needs to balance a trade-off between creating broad and sensitive searches (high sensitivity but low precision -> high workload) and high precision and restrictive searches (high precision but limited sensitivity -> low workload and more rapid synthesis). It is difficult to apply this balance in a consistent manner for all outcomes because the trade-off between sensitivity and precision will be different for each outcome and dependent on the overall amount of evidence for each research question.

On the practical side, one weakness is that only part of our workflow uses freely available software with a user-interface. The rest of our method, although available as a python scripts, requires data science or programming experience. However, this encourages the formation of an interdisciplinary team of researchers and drives a team science mentality which is important when tackling global health challenges. The need for human oversight may be seen as another limitation. Some of the methods, for example identifying sections via a rule-based approach, require further tailoring to individual research projects and cannot be used out-of-the-box. For the automated sentence classification, some human labelling of relevant sentences is needed. While human involvement does require resources, it also helps to reduce automation errors which leads to more streamlined processing and resource reductions downstream.

### Future research

For future research there is great promise in testing the value provided by LLMs, specifically large generative models for text processing. At the time of developing and applying the methodology described in this article, the context-length of leading LLMs was insufficient. GPT3.5, which was available at the time, had a context length of 16.000 tokens which corresponds to an average of

12.300 words.^22^ By context length we specifically refer to the amount of text processed by the LLM, including prompts, evaluation report text input, and for some LLMs including output length, too. On average, the evaluation report PDFs available to us had 57.843 words (± 27.913 standard deviation) which would have required an extensive amount of computations and multiple submissions for each report, repeated for each outcome. Additionally, these early LLMs were known to suffer from hallucinations, where the models created salient sounding, but factually incorrect text and it was beyond the scope of this project to perform manual checking for 64 outcomes across 631 evaluations. For this reason we decided to adopt the approach of more explainable classification on the sentence level first together with a methodology based on human-in-the loop.

Recent developments of LLM infrastructure led to increases of context length to 128- 200.000 tokens (98 – 153.000 words), decrease of hallucination (as claimed by model creators), as well as options to create structured outputs for multiple prompts being submitted at the same time.^23^ Using these recent models such as GPT-4o^24^ could further streamline the detection of information relevant to UNICEF’s strategic plan or SDG outcomes within evaluation full texts, but firstly needs to be tested using a well-designed and meaningful validation methodology.

## Conclusion

While text-mining and filtering methods are not expected to provide 100% complete results, they can be used to expedite the analysis of complex documents, such as evaluation reports. The methodology presented in this paper is most useful when rapidly analysing a large body of documents, focussing on breadth and accuracy rather than depth and sensitivity of results. By selecting relevant text via an existing summary section (expert-led) and then supplementing it with text-mining (AI-supported) we cut down the amount of irrelevant text presented to human data extractors. Then, by employing comprehensive and systematic search strategies to filter documents for each outcome in a human-in-the-loop system we aimed to boost transparency and reproducibility in the overall process. We provide our code for PDF-to-text, section processing, text-mining, and automatic creation of comprehensive adjacency-based tagging strategies within a python package. We hope this will encourage uptake of automation methods to support researchers interested in synthesising impact evaluations, reports, or grey literature in general.

Integrating natural language processing (NLP) to synthesize UNICEF evaluation reports (or similar) will necessitate significant digital infrastructure advancements. Key among these is the adoption of more structured evaluation reports and standardized templates that will facilitate enhanced machine readability. These changes will require the implementation of unified formatting and consistent terminologies to ensure that NLP algorithms can accurately interpret and process the content. Additionally, transitioning to digital-first documentation practices will support automated data extraction, analysis, and synthesis, enabling more efficient generation of insights from the vast corpus of evaluations. This evolution will enhance the ability to rapidly distil critical findings, trends, and lessons learned, fostering more effective decision-making and resource allocation within UNICEF.

## Statements and Declarations

### Author’s contributions

LS: Methodology, Data Curation, Formal Analysis, Software, Visualization, Writing – Original Draft Preparation

PA: Methodology, Data Curation, Investigation, Validation, Writing – Review & Editing HO: Methodology, Data Curation, Validation, Writing – Review & Editing

EM: Conceptualization, Methodology, Writing – Review & Editing KN: Methodology, Data Curation, Writing – Review & Editing UKH: Methodology, Writing – Review & Editing

ND: Methodology, Data Curation, Writing – Review & Editing

DC: Methodology, Funding Acquisition, Writing – Review & Editing

FC: Conceptualization, Methodology, Data Curation, Investigation, Funding Acquisition, Writing –

### Review & Editing

#### Ethical considerations

Not applicable: No human participants, human data or human tissue were studied in this work.

#### Consent to participate

Not applicable.

#### Consent for publication

Not applicable

## Acknowledgements

We like to thank Celeste Lebowitz and Zlata Bruckauf from UNICEF for their feedback and advice throughout this project.

## Data availability statement

All programming code for the automations described in this paper is available on GitHub: https://github.com/NIHRIO/EvaluationSynthesisMethods

The weights for the trained SPECTER model for UNICEF data are available here: https://drive.google.com/drive/folders/1-0VXJcY_GKBNq6-5GprPvwdnTc4Raud3?usp=sharing

The SWIFT-Review project is available as Appendix 3, it can be loaded and used using the free desktop application available here: https://www.sciome.com/swift-review/

World map visualisations of included evaluations for each goal area are available in Appendix 4.

Results from the evaluation synthesis are available on the UNICEF website: https://www.unicef.org/evaluation/evidence-unicef-achievements-children-synthesis-unicef-evaluations

## Declaration of conflicting interest

The author(s) declared no potential conflicts of interest with respect to the research, authorship, and/or publication of this article

## Funding statement

This project was funded by the UNICEF Evaluation Office. The UNICEF Evaluation Office operates independently within the organization, with a mandate to produce impartial and rigorous evidence that informs UNICEF’s policies, advocacy efforts, and programmes. For further details, please refer to the revised evaluation policy, available at https://www.unicef.org/executiveboard/revised-evaluation-policy-unicef-srs-2023. LS, PA, HO, DC, and FC were in part supported by the NIHR Innovation Observatory (National Institute for Health and Care Research (NIHR) [HSRIC-2016-10009/Innovation Observatory]). The views expressed are those of the author(s) and not necessarily those of the NIHR or the Department of Health and Social Care.

## Footnotes

1 https://docs.un.org/en/E/ICEF/2023/27 and https://www.unicef.org/evaluation/reports (last accessed 06/08/2024)

2 https://www.unicef.org/evaluation/reports#/?&gerosRating=(blank),Not%20Rated,Missing,Unsatisfactory,Fair,Satisfactory,Highly%20Satisfactory&yearofCompletion=2023,2022,2021,2020,2019,2018 (last accessed 06/08/2024)

3 https://developmentevidence.3ieimpact.org/ (last accessed 06/08/2024)

4 http://web.undp.org/evaluation/media-centre/blogs/aida-2.shtml (last accessed 06/08/2024)

5 https://policy-practice.oxfam.org/keyword/impact-evaluation/ (last accessed 06/08/2024)

6 https://www.unicef.org/evaluation/reports#/ (Last accessed 01/07/2024)

7 https://www.unicef.org/evaluation/evidence-unicef-achievements-children-synthesis-unicef-evaluations

8 https://www.unicef.org/evaluation/reports (last accessed 06/08/2024)

9 https://www.unicef.org/evaluation/documents/global-evaluation-reports-oversight-system-geros-handbook-and-summary-2017 (last accessed 06/08/2024)

10 https://pypdf.readthedocs.io/en/stable/

11 https://huggingface.co/google-bert/bert-base-uncased (last accessed 20/03/2025)

12 https://huggingface.co/allenai/scibert_scivocab_uncased (last accessed 20/03/2025)

13 https://huggingface.co/prajjwal1/bert-mini (last accessed 20/03/2025)

14 https://huggingface.co/docs/transformers/en/model_doc/xlnet#xlnet (last accessed 20/03/2025)

15 https://pypi.org/project/googletrans/ (last accessed 16/04/2024)

16 https://pypi.org/project/googletrans/

17 https://scikit-learn.org/stable/modules/generated/sklearn.metrics.classification_report.html (last accessed 19/03/2025)

18 https://www.un.org/en/about-us/un-system (last accessed 06/08/2024)

19 http://web.undp.org/evaluation/media-centre/blogs/aida-2.shtml and https://aida.undp.org/landing (last accessed 06/08/2024)

20 https://medium.com/sdg-counting/aida-artificial-intelligence-for-development-analytics-4a1a3d0f47e8 (Last accessed 19/03/2025)

21 https://www.sciome.com/swift-review/ (Last Accessed 25/07/2024)

22 https://platform.openai.com/docs/models/gpt-3.5-turbo (last accessed 20/03/2025)

23 https://platform.openai.com/docs/models (last accessed 20/03/2025)

24 https://platform.openai.com/docs/models/gpt-4o (last accessed 20/03/2025)

## Notes

### Competing Interest Statement

The authors have declared no competing interest.

### Summary of Updates

We integrated feedback, edited the abstract length, and expanded the Discussion and Future Works section.

## References

1. Cohan A, Feldman S, Beltagy I, et al. (2020). SPECTER: Document-level Representation Learning using Citation-informed Transformers

2. Devlin J, Chang MW, Lee K, et al. (2018). BERT: Pre-training of Deep Bidirectional Transformers for Language Understanding. arXiv preprint arXiv:1810.04805.

3. Howard BE, Phillips J, Miller K, et al. (2016). SWIFT-Review: a text-mining workbench for systematic review. Systematic Reviews, 5(1), 87. 10.1186/s13643-016-0263-z

4. Tanner R, Boender C, Vela P, et al. (2024). Are We Getting There? A synthesis of UN systemevaluations of SDG 5. New York: UN Women. https://www.unwomen.org/sites/default/files/2024-05/are-we-getting-there-a-synthesis-of-un-system-evaluations-of-sdg-5-annexes-en.pdf

5. UNICEF. (2022). UNICEF Strategic Plan 2022–2025: Renewed ambition towards 2030. New York: UNICEF. https://www.unicef.org/reports/unicef-strategic-plan-2022-2025

